# Improved pathogen identification in sepsis or septic shock by clinical metagenomic sequencing

**DOI:** 10.1101/2025.04.04.25324949

**Authors:** Thorsten Brenner, Sebastian O. Decker, Yevhen Vainshtein, Silke Grumaz, Mehdi Manoochehri, Manuel Feißt, Andrea Seidel-Glätzer, Mathias W. Pletz, Hendrik Bracht, Marc M. Berger, Kristina Fuest, Manfred Blobner, Friedhelm Bach, Onnen Moerer, Timo Brandenburger, Thomas Dimski, Klaudiusz Suchodolski, Ulrike Jäkel, Jana Zischkau, Helene Häberle, Peter Rosenberger, Tobias Schürholz, Simone Lindau, Stefan J. Schaller, Christian Putensen, Fabian Dusse, Sirak Petros, Max Gaasch, Christian Nusshag, Markus A. Weigand, Kai Sohn, German Society of Anaesthesiology and Intensive Care (GSAIC) Trials Group

## Abstract

**Objectives.:** Despite limited sensitivity and specificity, blood cultures (BCs) still represent the gold standard of diagnostic care in septic patients. We aimed to overcome current diagnostic limitations by unbiased next-generation sequencing (NGS) of circulating microbial cell-free DNA (mcfDNA) in plasma samples.

**Methods.:** We performed a prospective, observational, non-interventional, multicenter study (*Next GeneSiS-Trial*) to compare positivity rates for NGS-based identification of causative pathogens with BCs in patients suffering from sepsis or septic shock. An independent expert panel (n=3) retrospectively evaluated the plausibility of NGS-based findings and the potential for anti-infective treatment adaptations based on NGS results.

**Results.:** The positivity rate of NGS-based diagnostics (NGS+) for 491 septic patients was 70.5% compared to positive BCs (BC+) with 19.4% within the first three days after sepsis onset. NGS+ results were evaluated as plausible in 98.6% of cases by the expert panel. Based on the expertś recommendations, additional knowledge of NGS-based pathogen findings would have resulted in anti-infective treatment adaptations in 32.6% of all patients. Potentially inadequately treated NGS+/blood culture negative (BC-) patients showed worse outcomes.

**Conclusion.:** The integration of NGS-based pathogen diagnostics in sepsis has the potential to improve patientś outcomes as compared to a treatment strategy based on standard-of-care microbiological diagnostics alone.

## Introduction

### Pathophysiology and epidemiology of sepsis

Sepsis is defined as life-threatening organ dysfunction caused by a dysregulated host response to infection (1). With up to 50 million cases per year and an estimated global mortality rate of 11 million deaths per year, sepsis is responsible for nearly 20% of deaths worldwide (2). The same holds true for Germany, where sepsis accounts for up to 50% of hospital deaths and more than 20% of total deaths (3). Sepsis therefore represents one of the major global health threats, resulting in the WHO Resolution WHA 70.7 from May 29, 2017, titled ’Improving the prevention, diagnosis and clinical management of sepsis (4).

### Guideline-compliant sepsis care

Early identification of sepsis and timely implementation of measures within so-called ’sepsis care bundles’ (including administration of antimicrobials) is associated with significantly improved outcomes for both adult and pediatric patients (5–10). Accordingly, recent guidelines recommend an immediate administration of antimicrobials in adults with possible septic shock or a high likelihood for sepsis, ideally within 1 hour after recognition (11).

### Recent challenges in sepsis care

Particularly in critically ill patients with sepsis or septic shock, there is a high risk of inadequate empirical anti-infective therapy, which is unfortunately associated with significantly worse outcomes (12). However, the negative impact of an inadequate empirical therapy seems not to be limited to critically ill patients in intensive care units but also plays a relevant role in less severely ill hospitalized patients (13).

Accordingly, early identification of sepsis-causing pathogens is of utmost importance. Though, culture-based diagnostics represent the gold standard-of-care, the latter technique is associated with relevant diagnostic weaknesses (e.g. low sensitivity and specificity, prone to contaminations, long turn-around-times) (14).

### Metagenomic next-generation sequencing (NGS) for pathogen identification in sepsis

The detection of microbial circulating cell-free DNA (mcfDNA) by metagenomic next-generation sequencing (NGS) has shown to be a promising alternative. Following a highly stringent bench-to-bedside approach, a retrospective proof-of-concept study was published in first place, in which the detection of mcfDNA by a significance scoring system was shown to be applicable for the identification of causative pathogens in seven patients with sepsis or septic shock (15). These findings were reevaluated in a secondary analysis of a monocenter cohort of 48 patients with sepsis or septic shock, revealing a positivity of 71% with a plausibility of 96% for the NGS-based approach (16). Moreover, additional knowledge of these NGS findings would have led to a change towards an adequate anti-infective treatment regimen in more than half of the cases, as assessed by eight independent infectious disease or intensive care specialists (16). Although there is increasing evidence for (i) a significantly enhanced diagnostic performance as well as (ii) a possible improvement of patientś outcomes due to a more appropriate anti-infective treatment regimen, a prospective multicenter evaluation of the NGS-based approach in a large cohort of patients with sepsis or septic shock was still lacking (17–21). The *Next GeneSiS* (*<u>Next</u>*-*<u>Gene</u>*ration *<u>S</u>*equencing diagnostics of bacteremia in *<u>S</u>*epsis)-Trial was designed as a Germany-wide, multicenter, prospective, observational, non-interventional clinical study to assess detailed performance characteristics of the NGS-based approach and its potential impact for patientś anti-infective treatment regimen in a comprehensive cohort of patients with sepsis or septic shock (22).

## Materials and Methods

### Study design

The *Next GeneSiS-Trial* was a prospective, observational, non-interventional, multicenter, clinical study, which was conducted from 03/2019 to 09/2020 at medical and surgical intensive care units (ICUs) of 17 maximum care hospitals of the *German Society of Anaesthesiology and Intensive Care (GSAIC) Trials Group* (German Clinical Trials Register: DRKS00011911, ClinicalTrials.gov: NCT03356249) (22). Before the initiation of the study, the study protocol, patient information and informed consent, and all other required documents were submitted to the respective ethical review committees of all participating centers. The first positive ethical vote was given by the Ethics Committee of the Medical Faculty of Heidelberg (Trial Code No. S-084/2017). All study procedures were meant to ensure that all parties involved abide by the principles of Good Clinical Practice (GCP) and those stipulated in the Declaration of Helsinki. Patients with suspected or proven sepsis/septic shock according to the latest sepsis definitions (Sepsis-3) (1) with a diagnosis <24h were eligible for study inclusion. A summary of all inclusion and exclusion criteria is provided in **Supplemental Table 1**. Treatment of patients with sepsis or septic shock was performed according to the guidelines of the surviving sepsis campaign (SSC) applicable at that time (23). All participating study patients or their legal representatives signed written informed consent. Types of data collected are summarized in **Supplemental Methods**.

### Sample collection and standard-of-care microbiological analyses

Sample collection included two sets of blood cultures (BCs: 2x aerobic/ 2x anaerobic) at sepsis diagnosis and at 72h. In parallel, plasma samples for NGS-based measurements were obtained. Further blood samples for NGS-based measurements were collected whenever physicians ordered further BCs (2x aerobic / 2x anaerobic) within the first three days after sepsis diagnosis. BCs were considered positive for a patient when at least one bottle from the whole set of BC bottles showed microbial growth for a given day. Results of microbiological routine diagnostics in specimens different from blood (e.g. urine, tissue samples, wound swabs, bronchoalveolar lavage fluid (BALF), endotracheal aspirate) were considered for further analyses if they were obtained within ≤72 hours prior to or after the timepoints for NGS-based measurements.

Standard-of-care microbiological analyses of potential pathogens in the different specimens were performed according to the usual practice in each participating institution.

### NGS-based workflow

Plasma DNA was isolated either with a QiaSymphony (Qiagen, Hilden, Germany) as previously described (15) or with a QiaCube (Qiagen) using the QIAamp MinElute ccfDNA Kit according to the manufacturer’s instructions. When plasma volumes were below 1 ml, samples were supplemented up to 1 ml with sterile phosphate-buffered saline. For each batch of patient samples, one negative and one positive control were included. Quality control of isolated cfDNA was done using Qubit 3.0 Fluorometer (Thermo Fisher Scientific, Waltham, MA, USA) and Fragment Analyzer (Agilent Technologies, Santa Clara, CA, USA). Library preparation from 1 ng cfDNA was performed using the NEXTFLEX Cell Free-DNA Seq Kit Version 14.09 (Bioo Scientific, Austin, TX, USA) with a Biomek FXP liquid handling robot (Beckman Coulter, Brea, CA, USA). In short, end repair, adapter ligation and PCR were done according to the manufacturer’s protocol. Samples were sequenced using HiSeq2500 or NextSeq2000 in 100 base pairs single read mode and a sequencing depth of approximately 30 million reads per sample. Sequencing data analyses were performed using the Sepsis Indicating Quantifier (SIQ)-Score as described previously (15, 16, 22) and in **Supplemental Methods**. The datasets generated and/or analyzed during the current study are available from the corresponding author on reasonable request. Moreover, microbial sequencing data are available at the European Nucleotide Archive (ENA). ENA accession number for the Next GeneSiS-Trial sequencing data is: **PRJEB64401**

### Outcome analyses by expert panel evaluation

The clinical value of this NGS-based approach was assessed by a panel of three independent experts (i.e., specialists in infectious diseases and intensive care medicine not being associated with any of the participating study sites), retrospectively evaluating the plausibility and the therapeutic consequence of additional knowledge of the NGS-based test results. For all 491 patients that qualified for the expert panel evaluation (**Figure 1**), a patient report was generated, based on comprehensive clinical patient data extracted from the REDCap based electronic case report forms (eCRF) and SIQ-score analysis results (**Supplemental Figure 3**). These clinical patient data not only included microbiological data but also numerous other pieces of information regarding the medical history, clinical symptoms, clinical course, and laboratory values. The expert panel therefore attempted to simulate a situation that corresponds to that of the clinician at the patient’s bedside; the only difference was that the reviewers could additionally refer to the data from NGS diagnostics. To evaluate the clinical relevance of the NGS-based approach, reports were presented in combination with a questionnaire on an online survey platform of the Fraunhofer Society to the clinical experts. A detailed description of the questionnaire is outlined in **Supplemental Figure 4**. In the case assessment regarding a change in the therapy regimen, the experts were explicitly asked to consider only recommendations for changes based on the diagnostic added value of NGS diagnostics. General therapy changes within the framework of Antibiotic Stewardship (ABS) principles that would have arisen independently of NGS diagnostics should explicitly not be taken into account.

**Figure 1.**
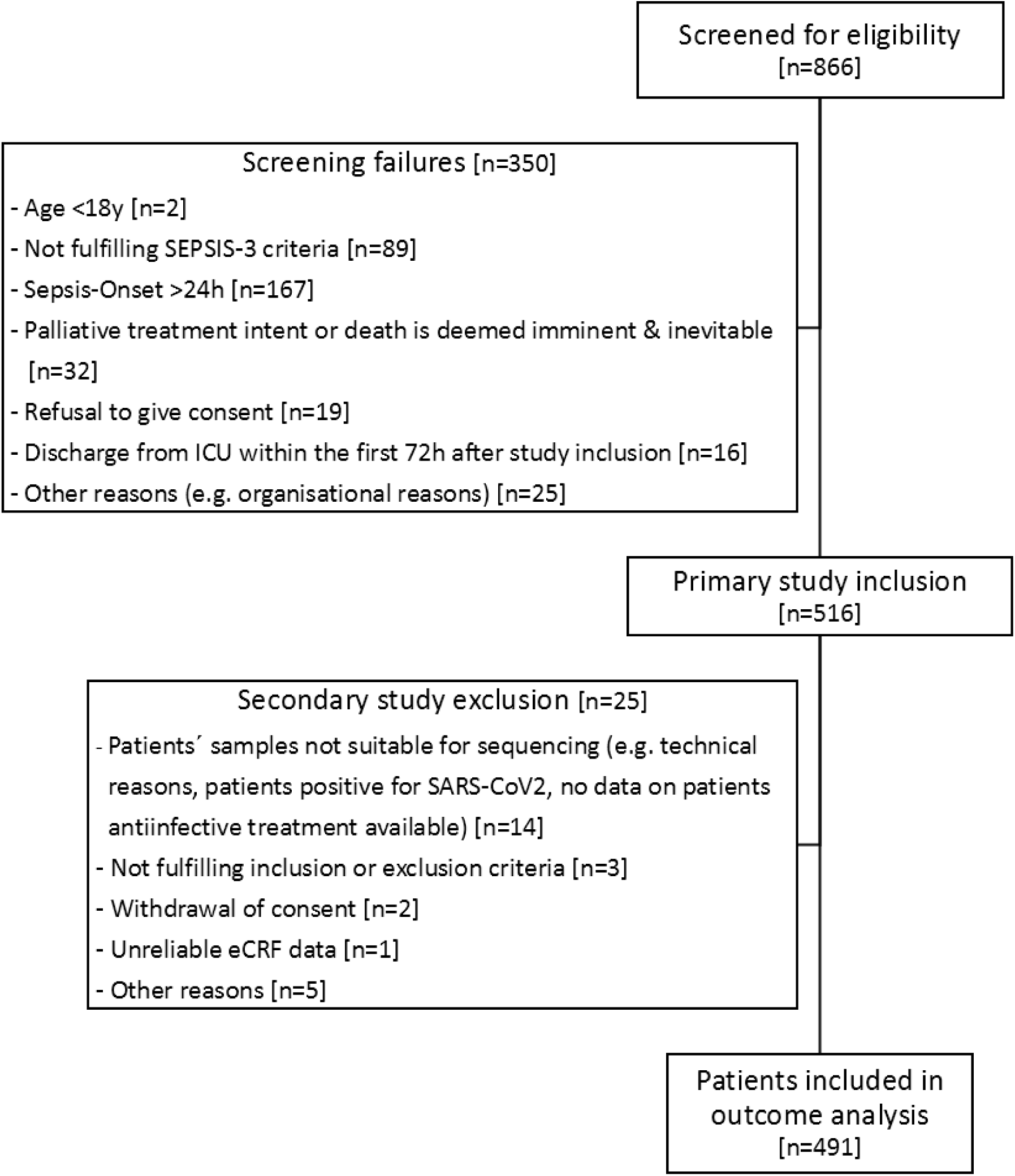
Study flow chart. This figure illustrates the study design of the *Next GeneSiS-Trial*. A total of 866 patients were screened for eligibility. Of these, 350 patients had to be considered as screening failures and could therefore not be considered for further study participation, resulting in 516 patients with primary study inclusion. After the secondary study exclusion of 25 patients, 491 patients could be included in the outcome analysis. <u>Abbreviations:</u> eCRF, electronic case report form; SARS-CoV2, severe acute respiratory syndrome coronavirus type 2

### Statistical methods

The *Next GeneSiS-Trial* was performed in terms of an exploratory pilot study and was therefore not statistically powered in a formal sample size calculation procedure. It was estimated, that approximately 500 patients had to be enrolled to enable a reasonable assessment of the performance of the NGS-based approach when compared with standard-of-care microbiological diagnostics. For an evaluation of the NGS-performance, results obtained with the NGS technology for each sample were then compared with those obtained using conventional microbiology methods for the same sample. Agreement and concordance were assessed using a McNemar test and Cohen κ. All percentages and confidence intervals (Cis) for proportions were calculated using the exact method and rounded to the nearest percentage. The clinical value of the NGS-based approach was estimated by a panel of three independent clinical specialists not associated with the participating study sites by the use of the above-mentioned questionnaire. Analyses of the reviewers’ independent responses were performed using a majority rule such that two of three responses for a given patient determined the outcome for that patient. Comparison between subgroups was performed by unadjusted χ2 tests for categorical data and methods of variance analysis for continuous data. Analyses were performed with all available data, so that no imputation of missing values was performed. All statistical tests were performed using SAS (SAS Institute, Cary, NC). A p-value of less than 0.05 was considered statistically significant in a descriptive sense. In addition, graphs are presented wherever possible.

## Results

### Study cohort and state-of-the-art microbiological diagnostics

In total, 866 patients with sepsis or septic shock were screened for eligibility, of which 350 patients had to be excluded (**Figure 1**) in terms of screening failures and 25 patients could not be considered for outcome analysis (e.g. patients’ samples not suitable for sequencing, withdrawal of consent). Patientś mean age was 67 years with a predominance of males (67.4%; **Table 1**). Participating patients revealed a small number of comorbidities as assessed by a mean Charlson Comorbidity Index (CCI) of 2.9±2.4 and suffered from immunosuppressive host factors in 8.9%. Septic shock was present in 305 (70.9%) of all included patients with a mean SOFA score of 9.1±3.4. The mode of acquisition was evenly distributed between outpatient (51.9%) and nosocomial (48.1%). The primary septic focus was the abdomen (62.1%), followed by the lung (36.0%) and the genitourinary tract (23.6%). In total, 439 infectious source control measures were performed in 312 (63.5%) patients, predominantly surgical (71.5%). Anti-infectives were given for a mean duration of 11.7±8.1 days with an average number of 2.96 anti-infectives per patient (**Supplemental Table 2**). The anti-infective treatment regimen primarily consisted of antibiotics (88.6%) with beta lactam antibiotics, such as penicillins (28.0%) or carbapenems (19.1%) as the primary substance groups. In contrast, antifungals (9.0%) and virostatic agents (2.3%) were used less frequently. The overall 28-day mortality was 26.7% with a mean length of ICU/ intermediate care (IMC) unit stay of 11.4±8.9 days. A need for mechanical ventilation as well as kidney replacement therapy (KRT) was present in 73.9% and 26.1%, respectively. The length of mechanical ventilation as well as KRT was 8.7±8.5 days and 9.3±8.4 days. Within a timeframe of three days before sepsis onset up to six days following study inclusion, a total number of 8.167 microbiological samples were analyzed with an average of 16.6±7.2 microbiological samples per patient (**Supplemental Table 3**) with BCs being the most frequently used material for microbiological analyses (5.536 BC bottles; 67.8%). The rest of the samples used for microbiological analyses are displayed in more detail in **Supplemental Table 3**.

**Table 1.** Patientś demographics, clinical data and outcome characteristics.

| **Demographic Data** |  |
| --- | --- |
| Age [y] | 67.0±14.1 |
| Male sex | 331 (67.4%) |
| **Comorbidities** |  |
| Charlson Comorbidity Index (CCI) | 2.9±2.4 |
| **Immune status** |  |
| Immunosuppressed | 44 (8.9%) |
| **Source of infection** |  |
| Outpatient | 255 (51.9%) |
| Nosocomial | 236 (48.1%) |
| **Infection sites (multiple allocations possible)** |  |
| Pulmonary / upper airway / thoracic | 177 (36.0%) |
| Intraabdominal / biliary / gastrointestinal tract | 305 (62.1%) |
| Genitourinary tract | 116 (23.6%) |
| Surgical site infection | 26 (5.3%) |
| Bone, joint and soft tissue infections | 36 (7.3) |
| Others | 81 (16.5%) |
| Missing / Unknown | 22 (4.5%) |
| **SEPSIS-3 criteria (at study inclusion)** |  |
| Infection (suspected or proven) | 491 (100.0%) |
| SOFA score | 9.1±3.4 |
| Lactate [mg/dl] | 34.6±34.2 |
| Septic shock | 305 (70.9%) |
| **Infectious disease biomarkers** (within 24h following study inclusion) |  |
| Leucocyte count [$10^3/\mu\text{l}$] | 15.7±9.6 |
| C-reactive protein [mg/dl] | 20.4±13.2 |
| Procalcitonin [ng/ml] | 29.9±67.4 |
| **Infectious source control** (up to 6 days following study inclusion) |  |
| No. of patients (with infectious source control measures) | 312 (63.5%) |
| No. of measures in these patients | 439 |
| Type of infectious source control measures: |  |
| - Surgical | 314 (71.5%) |
| - Interventional | 85 (19.4%) |
| - Others | 40 (9.1%) |
| **Clinical outcome data** (at or within 28 days following study inclusion) |  |
| Mortality | 131 (26.7%) |
| Length of ICU/IMC stay [days] | 11.4±8.9 |
| Need for mechanical ventilation | 363 (73.9%) |
| Length of mechanical ventilation [days] | 8.7±8.5 |
| Length of anti-infective treatment [days] | 11.7±8.1 |
| Need for KRT | 128 (26.1%) |
| Length of KRT [days] | 9.3±8.4 |
| Data are presented by number (%) or mean ± standard deviation. |  |
| <u>Abbreviations:</u> CCI, Charlson Comorbidity Index; SOFA, Sequential Organ Failure Assessment, ICU, intensive care unit, IMC, intermediate care; KRT, kidney replacement therapy |  |

### Performance characteristics of BC and NGS-diagnostics

For a direct comparison of BC-results with NGS, same-day blood samples were acquired for each procedure at diagnosis of sepsis and 72h later. In total, 489 sets of BCs (each set consisting of at least four BC bottles) were cultured at the onset of sepsis and 406 additional sets 72h later. For NGS-based analyses, blood was drawn from patients at onset (491 samples) and 72h later (414 samples). Positivity for NGS-based pathogen identification was 76.2% at diagnosis of sepsis and 63.8% 72 h later. The average combined positivity rate for both time points was 70.5%. (**Figure 2a**). In contrast, positivity for BCs was significantly lower with 28.4% positive BC sets at sepsis diagnosis and 8.6% at 72 h resulting in a combined positivity rate of 19.4% (**Figure 2a**) and a three times lower sensitivity than that of NGS. Moreover, experts confirmed that 98.6% of all NGS positive results combined for both time points were plausible (97.9% plausibility rate at sepsis diagnosis, 99.6% 72 h later) (**Figure 2b**). Even NGS negative results were judged plausible in 92.5% of all cases at both time points (**Figure 2b**). A concordance of NGS to blood culture results was found for 45.7% of samples at both time points combined (**Supplemental Tables 5-7**). The most frequent species detected by BC and NGS was found to be *E. coli* (**Figures 3a**, **3b, 3d**). Additionally, NGS detected more species per sample (mean: 3.22 species/ NGS-positive sample) compared to BCs (mean: 1.11 species/positive BC bottle) indicative of polymicrobial sepsis. Moreover, bona fide contaminants were more effectively removed by NGS-based diagnostics using SIQ-score as compared to BCs, with *S. epidermidis* representing the most frequently detected species in the BC positive/NGS negative (BC+/NGS-) subgroup analysis (**Figure 3c; Supplemental Table 4**).

**Figure 2.**
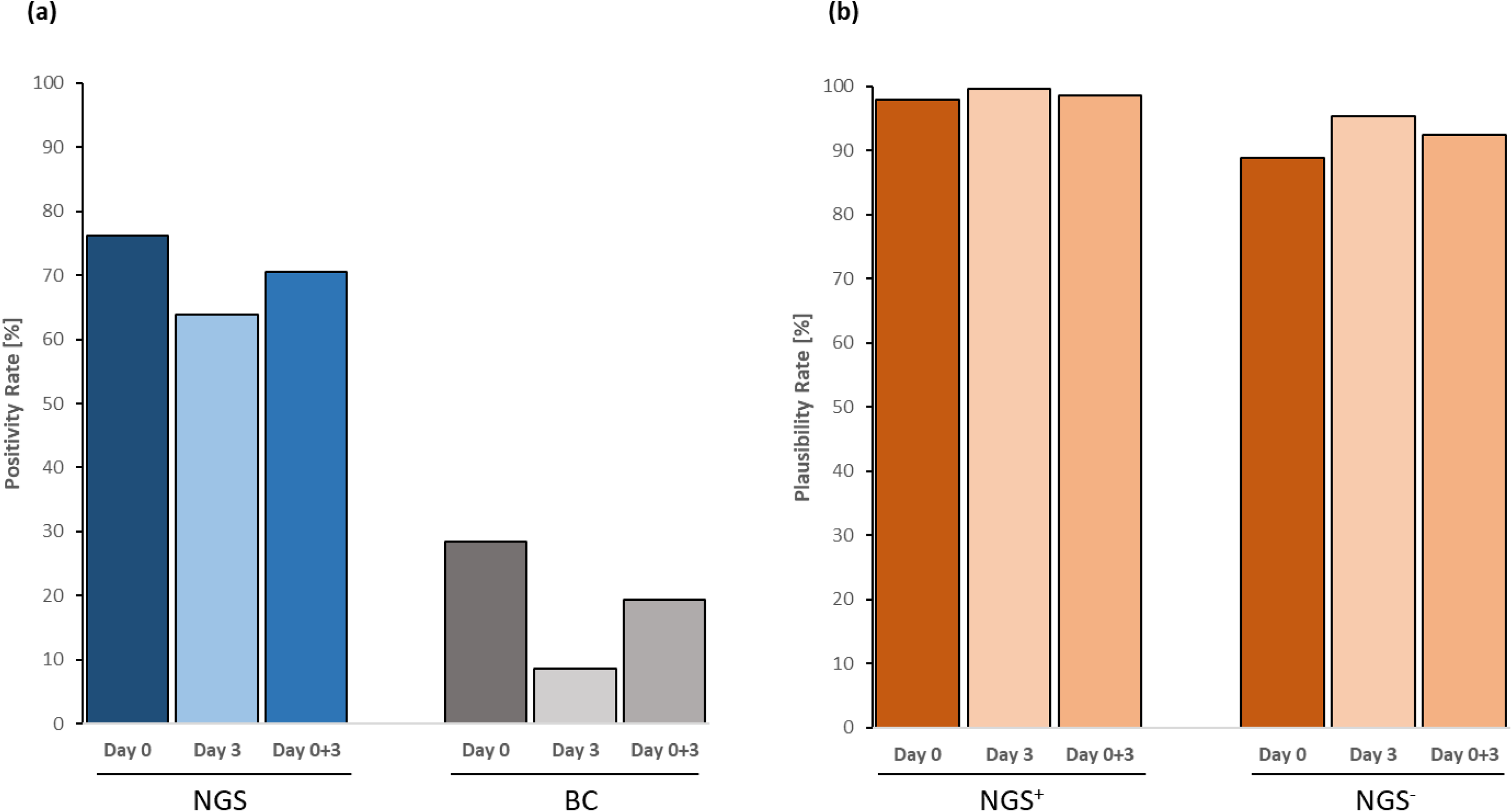
Positivity and plausibility rates of next-generation sequencing (NGS) and blood culture (BC) diagnostics. **(a)** Positivity rates of NGS-based diagnostics and BC at sepsis onset (day 0), 72 h after sepsis onset (day 3) as well as at both time points combined (day 0-3). **(b)** Plausibility rates of NGS-based diagnostics are presented separately for NGS-positive (NGS +) and NGS-negative (NGS -) results at sepsis onset (day 0), three days after sepsis onset (day 3) as well as at both time-points combined (day 0-3). <u>Abbreviations:</u> BC, blood culture; NGS, next-generation sequencing

**Figure 3.**
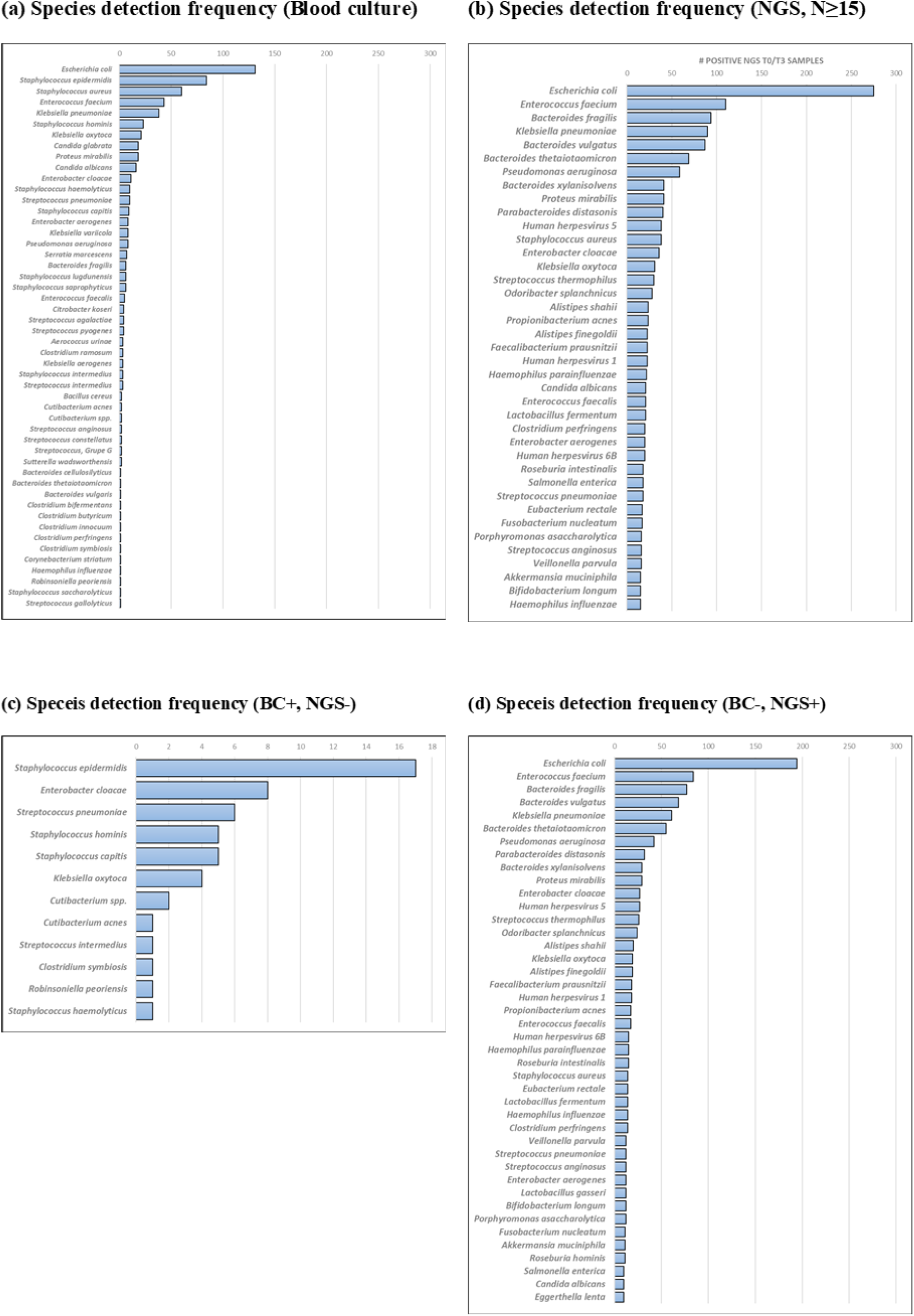
Frequencies of detected sepsis-causatives by blood culture (BC) as well as next-generation sequencing (NGS). Species detected by **(a.)** BC and **(b.)** NGS over the study time points. The most frequently detected species by both techniques was shown to be *E. coli*, with an additional overlap in other species. However, a proportion of species is only observed in either one of the methods. (c) Species detected by BC but not by mNGS (BC+ / NGS−). The most frequently detected species is *Staphylococcus epidermidis*, consistent with its common role as a contaminant in blood culture. (d) Species detected by mNGS but not by BC (BC− / NGS+). <u>Abbreviations:</u> BC, blood culture; NGS, next-generation sequencing

### Clinical value of the NGS-based approach

Based on NGS results, the independent expert panel recommended a change in the individual treatment regimen in 160 patients (32.6%) of the whole cohort of septic patients. For 105 patients, experts proposed a de-escalation of the anti-infective treatment regimen, whereas an escalation of therapy was suggested in 49 patients (**Figure 4**). Among those, a combination of both treatment adaptations was recommended in nine patients. In 79 of 227 patients (34.8%) with an empiric treatment failure within the first three days after sepsis onset (as defined by death of the patient <u>or</u> lack of improvement of the patient’s clinical condition (in terms of an inadequate decrease of SOFA score) <u>or</u> persistent high procalcitonin levels), a NGS-based change in the individual treatment regimen was recommended by the post-hoc expert panel, which was significantly less frequently considered in patients without an empiric treatment failure (66 of 249 patients; 26.5% / p=0.0495*).

**Figure 4.**
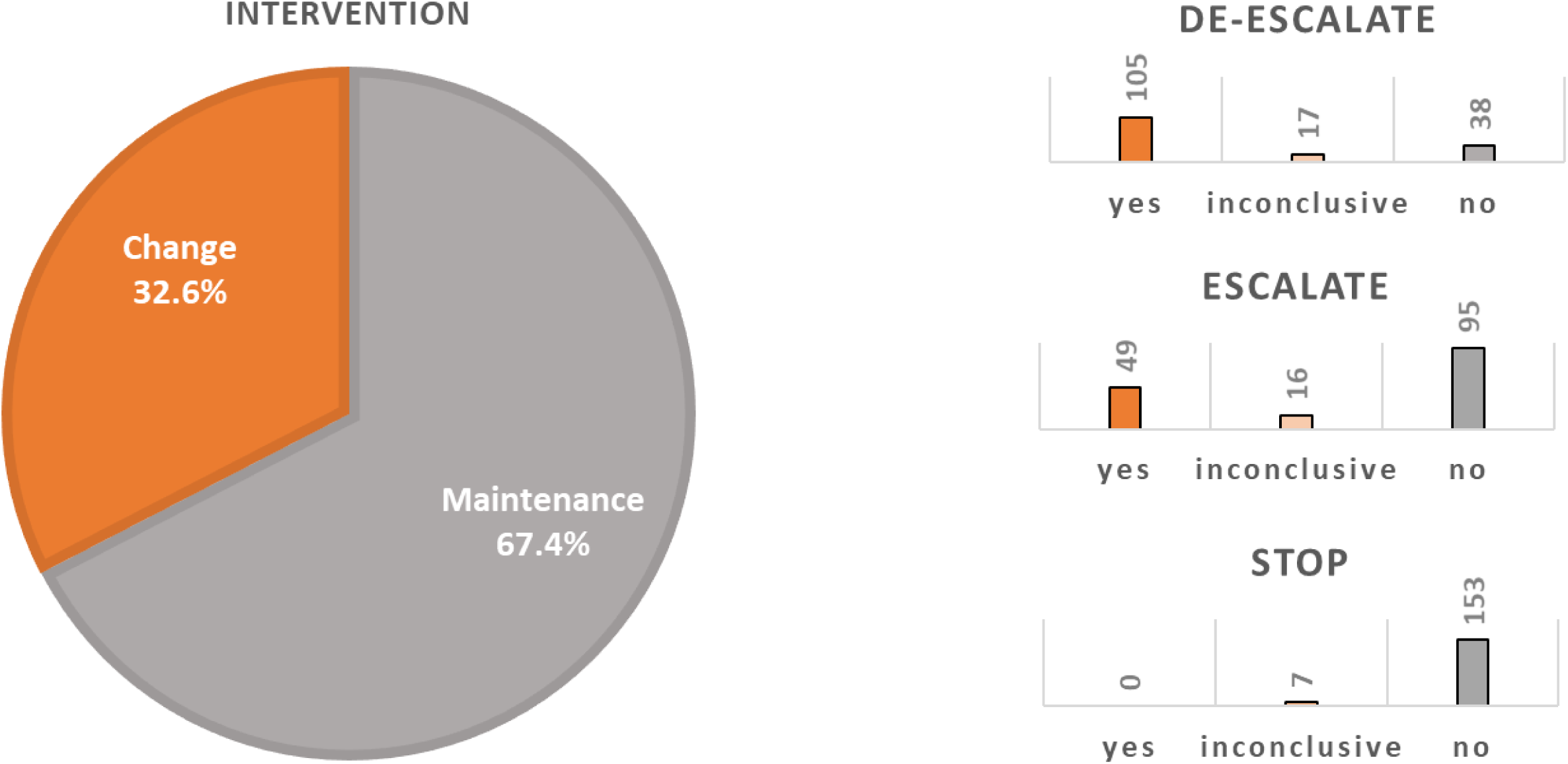
Expert panel-based recommendations for patientś anti-infective treatment regimen. Expert evaluation of the impact of next-generation sequencing (NGS) results on the antimicrobial treatment regimen. Results from the majority vote regarding a hypothetical anti-infective treatment intervention based on the NGS findings, resulted in a change recommendation in 32.6% of all patients (left-sided pie chart). The resulting change recommendations (de-escalate, escalate, stop) are presented in detail in the right-sided bar charts. For clear “yes” or “no” statements at least two of the three experts had to recommend (1.) a change of the anti-infective treatment based on the NGS findings (see Supplemental Figure 4 → question 2 (q2) with the answer “no”) and (2.) a treatment change pointing towards the same direction (see Supplemental Figure 4 → questions 3 (q3) a-c with at least two experts answering either “yes” or “no”). <u>Abbreviations:</u> NGS, next-generation sequencing

Remarkably, lengths of antifungal and antiviral treatments as well as the duration of KRT were significantly different between the treatment change subgroups. These differences were particularly evident in patients for whom a de-escalation of the anti-infective treatment regimen was recommended in both, the complete cohort as well as in surviving septic patients (**Table 2a, 2b**). Considering the subgroup of NGS(+)/BC(-) patients (n=205), patients with recommendations for anti-infective treatment changes (n=93) experienced significantly worse outcomes (including lengths of ICU/IMC stay, mechanical ventilation, KRT, antibiotic as well as antiviral treatments) as compared to those with a recommendation for maintaining the treatment strategy (**Table 3a, 3b, Supplemental Table 8a, 8b**).

**Table 2a.**
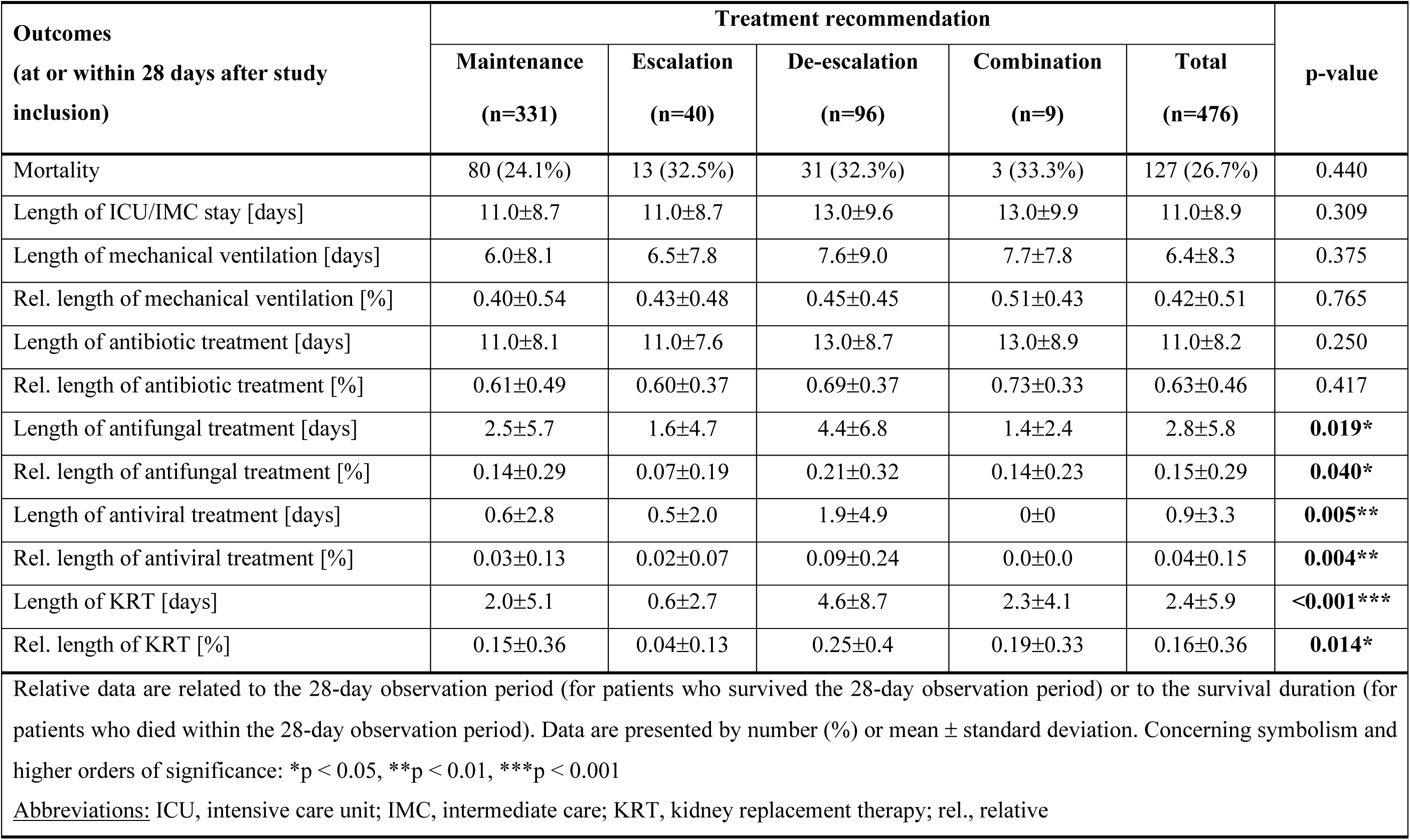
Treatment recommendations with absolute and relative clinical outcome characteristics in the whole cohort of septic patients.

**Table 2b.**
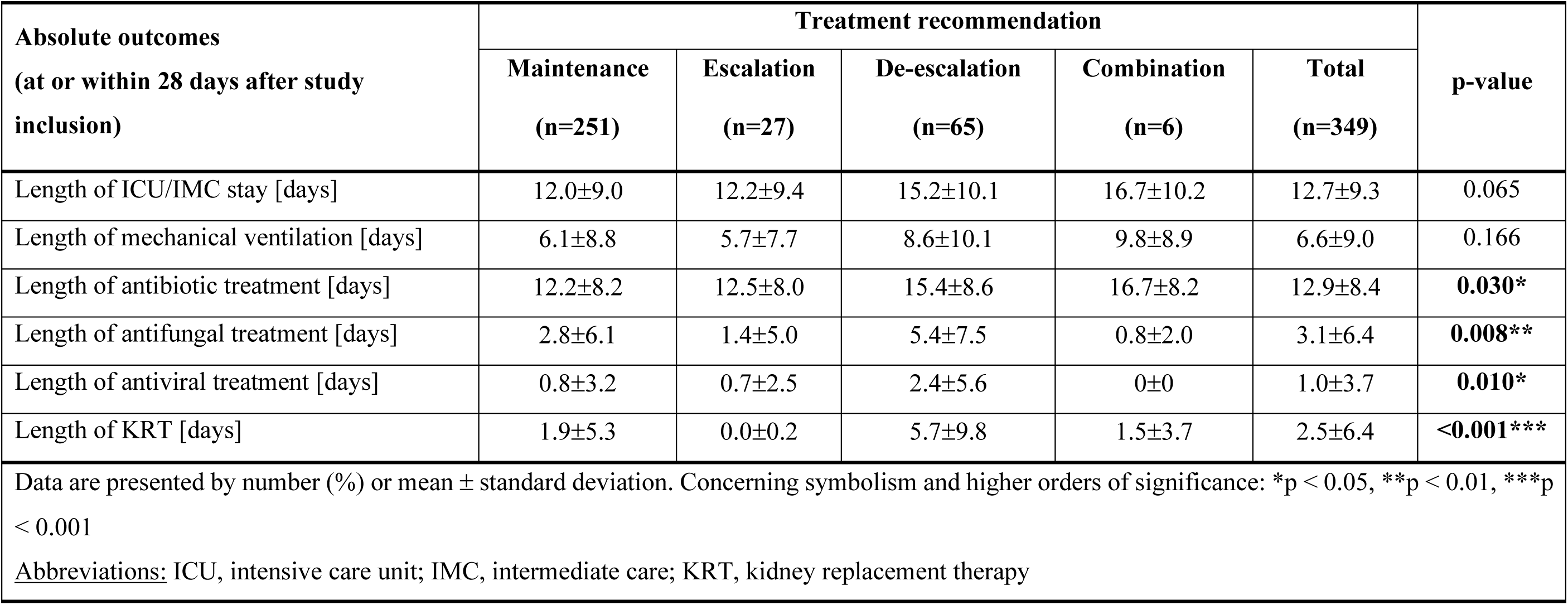
Treatment recommendations and absolute clinical outcome characteristics in surviving septic patients.

| Absolute outcomes<br>(at or within 28 days after study<br>inclusion) | Treatment recommendation |  |  |  |  | p-value |
| --- | --- | --- | --- | --- | --- | --- |
|  | Maintenance<br>(n=251) | Escalation<br>(n=27) | De-escalation<br>(n=65) | Combination<br>(n=6) | Total<br>(n=349) |  |
| Length of ICU/IMC stay [days] | 12.0±9.0 | 12.2±9.4 | 15.2±10.1 | 16.7±10.2 | 12.7±9.3 | 0.065 |
| Length of mechanical ventilation [days] | 6.1±8.8 | 5.7±7.7 | 8.6±10.1 | 9.8±8.9 | 6.6±9.0 | 0.166 |
| Length of antibiotic treatment [days] | 12.2±8.2 | 12.5±8.0 | 15.4±8.6 | 16.7±8.2 | 12.9±8.4 | **0.030\*** |
| Length of antifungal treatment [days] | 2.8±6.1 | 1.4±5.0 | 5.4±7.5 | 0.8±2.0 | 3.1±6.4 | **0.008\*\*** |
| Length of antiviral treatment [days] | 0.8±3.2 | 0.7±2.5 | 2.4±5.6 | 0±0 | 1.0±3.7 | **0.010\*** |
| Length of KRT [days] | 1.9±5.3 | 0.0±0.2 | 5.7±9.8 | 1.5±3.7 | 2.5±6.4 | **<0.001\*\*\*** |
| Data are presented by number (%) or mean ± standard deviation. Concerning symbolism and higher orders of significance: *p < 0.05, **p < 0.01, ***p < 0.001 |  |  |  |  |  |  |
| <u>Abbreviations:</u> ICU, intensive care unit; IMC, intermediate care; KRT, kidney replacement therapy |  |  |  |  |  |  |

**Table 3a.** Treatment recommendations and clinical outcome characteristics in a NGS(+) and BC(-) subgroup of septic patients.

| Outcomes<br>(at or within 28 days after study inclusion) | Treatment recommendation |  |  | p-value |
| --- | --- | --- | --- | --- |
|  | Maintenance<br>(n=112) | Change<br>(n=93) | Total<br>(n=205) |  |
| Mortality | 17 (15.2%) | 21 (22.6%) | 38 (18.5%) | 0.175 |
| Length of ICU/IMC stay [days] | 11.7±8.4 | 14.5±9.0 | 13.0±8.8 | **0.026\*** |
| Rel. length of ICU/IMC stay [days] | 0.54±0.41 | 0.64±0.37 | 0.59±0.39 | 0.084 |
| Length of mechanical ventilation [days] | 5.8±7.3 | 8.6±9.2 | 7.1±8.3 | **0.018\*** |
| Rel. length of mechanical ventilation [days] | 0.32±0.40 | 0.39±0.39 | 0.35±0.40 | 0.174 |
| Length of antibiotic treatment [days] | 11.7±7.4 | 14.0±8.1 | 12.7±7.8 | **0.038\*** |
| Rel. length of antibiotic treatment [days] | 0.53±0.36 | 0.61±0.32 | 0.57±0.34 | 0.123 |
| Length of antifungal treatment [days] | 3.1±6.6 | 4.4±6.8 | 3.7±6.7 | 0.169 |
| Rel. length of antifungal treatment [days] | 0.13±0.28 | 0.19±0.29 | 0.16±0.29 | 0.187 |
| Length of antiviral treatment [days] | 0.6±2.2 | 1.8±4.8 | 1.1±3.7 | **0.026\*** |
| Rel. length of antiviral treatment [days] | 0.02±0.08 | 0.07±0.20 | 0.05±0.15 | **0.017\*** |
| Length of KRT [days] | 1.7±4.8 | 3.9±8.0 | 2.7±6.5 | **0.021\*** |
| Rel. length of KRT [days] | 0.09±0.22 | 0.17±0.32 | 0.12±0.27 | **0.035\*** |
| Relative data are related to the 28-day observation period (for patients who survived the 28-day observation period) or to the survival duration (for patients who died within the 28-day observation period). Data are presented by mean ± standard deviation. Concerning symbolism and higher orders of significance: *p < 0.05<br><u>Abbreviations:</u> BC, blood culture; ICU, intensive care unit; IMC, intermediate care; KRT, kidney replacement therapy; NGS, next-generation sequencing; rel. relative |  |  |  |  |

**Table 3b.** Treatment recommendations and absolute clinical outcome characteristics in a NGS(+) and BC(-) subgroup of surviving septic patients.

| Absolute outcomes<br>(at or within 28 days after study inclusion) | Treatment recommendation |  |  | p-value |
| --- | --- | --- | --- | --- |
|  | Maintenance<br>(n=95) | Change<br>(n=72) | Total<br>(n=167) |  |
| Length of ICU/IMC stay [days] | 12.2±8.9 | 15.3±9.6 | 13.6±9.3 | **0.037\*** |
| Length of mechanical ventilation [days] | 5.6±7.7 | 8.7±9.6 | 6.9±8.7 | **0.029\*** |
| Length of antibiotic treatment [days] | 12.2±7.7 | 14.9±8.4 | 13.4±8.1 | **0.034\*** |
| Length of antifungal treatment [days] | 3.3±7.0 | 4.6±7.3 | 3.8±7.1 | 0.241 |
| Length of antiviral treatment [days] | 0.7±2.4 | 2.0±5.1 | 1.2±3.9 | **0.044\*** |
| Length of KRT [days] | 1.7±5.0 | 3.9±8.5 | 2.6±6.8 | 0.054 |
| Data are presented by mean ± standard deviation. Concerning symbolism and higher orders of significance: *p < 0.05 |  |  |  |  |
| <u>Abbreviations:</u> BC, blood culture; ICU, intensive care unit; IMC, intermediate care; KRT, kidney replacement therapy; NGS, next-generation sequencing |  |  |  |  |

## Discussion

With nearly 500 patients from 17 study sites the *NextGeneSiS-Trial* represents one of the most comprehensive, prospective investigations to robustly benchmark unbiased metagenomic diagnostics based on high-throughput sequencing of mcfDNA in septic plasma samples in comparison to BC-based standard-of-care diagnostics. Two other studies with at least 200 samples have been published for the evaluation of NGS-based diagnostics; however, none of both combined prospective sampling with clinical outcome expert evaluations (24, 25). In addition, for technical validation of performance of mcfDNA sequencing diagnostics a cohort of 350 septic patients has been analyzed, confirming superior performance compared to standard-of-care BCs, but also lacking any expert outcome evaluations (26). In contrast, our study included an in-depth review of NGS-based diagnostic findings concerning plausibility as well as their impact on potential treatment changes as assessed by an independent expert panel. Expert evaluations facilitated the stratification of patient groups based on the adequacy of underlying anti-infective treatment regimens in order to determine potential outcome differences for adequately versus inadequately treated patients.

BCs currently represent the gold standard of diagnostic care for pathogen detection in bloodstream infections including sepsis/septic shock (11). However, although two sets of BC bottles were taken per time-point for aerobic and anaerobic cultures, a combined positivity rate of only 19.4 % for BCs at onset and 72 hours later was rather low, which is in good agreement with published data (27, 28). Considering that a significant proportion of BC bottles revealed bona fide contaminants including coagulase-negative staphylococci (CoNS, e.g. *S. epidermidis* (second most abundant pathogen in BCs) or *S. hominis*), the actual number of BC bottles containing relevant pathogens is expected to be even lower. In contrast, the positivity rate for NGS-based diagnostics for both time-points together was 70.5% excluding bona fide contaminants by statistical relevance scoring using the SIQ score. This was more than three times higher than BC diagnostics, confirming earlier findings that NGS of mcfDNA represents a significantly more sensitive approach (16–21). This was also verified at the species level since *E. coli* was the most frequently detected pathogen by both methods. However, *E. coli* was detected more than three times more frequently by NGS as compared to BCs with respect to absolute numbers. In addition, NGS-based diagnostics detected on average three times more species per NGS(+) sample (mean 3.22) as compared to BC diagnostics (mean 1.11 per BC) demonstrating better suitability for the detection of polymicrobial infections in septic patients. Furthermore, NGS was able to detect anaerobic fermenting bacteria very frequently (e.g. *Bacteroides* spp.), which were almost absent in conventional BC. Difficult-to-diagnose and rare infections including fungal infections (e.g. candidemia, invasive aspergillosis) could also be detected by unbiased NGS-based diagnostics within the *NextGeneSiS-Trial*.

While our results demonstrate that NGS detects pathogens in a significantly higher proportion of cases compared to blood cultures (BCs), it is important to put these findings into context: The higher sensitivity of NGS, particularly in cases where BC results are negative, may reflect its ability to detect pathogens that are non-culturable, present at low abundance, or fastidious organisms that require specific growth conditions. However, we acknowledge that although we made considerable efforts to validate our findings by experts (NGS positive (NGS+) as well as NGS negative (NGS-) results were judged plausible in more than 98% and 92% of cases, respectively) the possibility that few of the NGS-positive BC-negative results might also represent false positives. These may arise due to the high sensitivity of NGS detecting DNA from contaminations during sample handling, for example. Further studies are needed to confirm the clinical relevance of NGS-positive findings, particularly in cases where BC results are negative.

Regarding dynamics, mcfDNA was also shown to exhibit a relatively short half-life of only few hours in a cecal ligation and puncture (CLP)-based animal model of sepsis (29). In this study bacterial NGS signals for *E. coli*, *B. vulgatus* and *Enterococcus* spp. almost completely vanished within 12-24h as soon as the underlying infectious source was under control. Therefore, mcfDNA represents a highly precise biomarker that only occurs as long as the infectious focus is present. In addition, NGS-based diagnostics only require low sample volumes of 1 ml or less, making it more suitable than microbial cultures especially in pediatric or neonatal sepsis where sample volumes are limited (30, 31).

Adequate antimicrobial therapy is crucial for the treatment success in septic patients (5–10). Vice versa, inadequate treatment is associated with worse outcome (32). Consequently, precise pathogen detection is of utmost importance in order to switch from an empiric to a more effective targeted antimicrobial treatment strategy as early as possible. Remarkably, the expert panel of the *NextGeneSiS-Trial* suggested an adaptation of the treatment regimen in one-third of cases to achieve more adequacy. This is in good agreement with a previous monocentric study, where treatment changes were recommended in approximately half of the cases (16). Among patients of the *Next GeneSiS-Trial* with a potentially inadequate anti-infective treatment regimen, experts recommended escalations as well as de-escalations of antimicrobials in approximately one-third and two-third of cases, respectively. The predominance of NGS-based recommendations for de-escalation of the existing anti-infective therapy might be due to the low sensitivity, sometimes long latency until results are communicated, and high susceptibility to contamination associated with (blood) culture diagnostics (14, 33). And although NGS diagnostics generally exhibit very high sensitivity for microbial nucleic acids, the SIQ score-based approach (which, as is well known, includes findings from healthy individuals as well as surgical controls) minimizes the risk of a high rate of false-positive results (in terms of contamination). Strikingly, in none of the cases a complete stop of empiric therapy was recommended. However, individual experts considered a termination of the antimicrobial treatment based on NGS results in seven cases. This reservation might be due to the fact, that NGS-based diagnostics are not yet widely established in clinical routine to support such far-reaching clinical decisions. It is interesting that those patients for whom the experts recommended a de-escalation of the anti-infective treatment regimen revealed a prolonged need for KRT. This is in line with previously published data, associating high concentrations or certain combinations of antimicrobials (piperacillin/tazobactam plus vancomycin) with worse outcomes (34, 35). Most interestingly, outcome differences were most pronounced in adequately versus potentially inadequately treated patients with positive NGS (NGS+) and concurrent negative BC (BC-) results. These patients differed in ICU length of stay as well as the duration of ventilation, potentially affecting long-term outcomes and socioeconomic burdens. To avoid any sources of bias and to guarantee reliable study results, the *NextGeneSiS-Trial* followed a highly stringent approach (e.g. multicentric study design, evaluation by three independent clinical specialists using a majority rule) providing evidence for worse outcomes due to insufficient standard-of-care diagnostics. However, the here presented investigation cannot demonstrate a causal relationship due to its non-interventional study design. This limitation. can only be overcome by an interventional randomized trial, in which NGS-based diagnostics are provided in real-time to be used for anti-infective therapy guidance at the bedside. Such an interventional multi-center study entitled *DigiSep-Trial* is on its way, assessing the outcome relevance of a holistic diagnostic approach (consisting of a combination of NGS- and culture-based diagnostics) as compared to culture-based standard diagnostics alone (36).

## Supporting information

Supplementary for Brenner

## Data Availability

The datasets generated and/or analyzed during the current study are available from the corresponding author on reasonable request. Moreover, microbial sequencing data are available at the European Nucleotide Archive (ENA). ENA accession number for the Next GeneSiS-Trial sequencing data is: PRJEB64401

## Acknowledgements

We thank all patients, referring physicians, and study nurses who submitted samples. Moreover, we would like to acknowledge Wolfgang Krüger, MD, Christian Lanckohr, MD and Stefan Hagel, MD for their valuable participation in the expert panel.

## Funding

This study received a financial grant from Dietmar Hopp Stiftung, Opelstraße 28, 68789 St. Leon-Rot, Germany and internal funding from Fraunhofer.

### Authors’ contributions

Thorsten Brenner and Kai Sohn each made substantial contributions to the design of the work and drafted the manuscript. Manuel Feißt was responsible for data management and statistical analyses. Andrea Seidel-Glätzer was responsible for project management and clinical monitoring. Silke Grumaz, Mehdi Manoochehri, Karolina Glanz and Yevhen Vainshtein were responsible for NGS-based diagnostics as well as bioinformatic analyses. Sebastian O. Decker, Mathias W. Pletz and Markus A. Weigand made substantial contributions to the conception of the work and revised the manuscript critically. The rest of the authors contributed substantially to the recruitment of study participants, provision of biosamples, collection of clinical data and critical revision of the manuscript. All authors revised the manuscript and have given final approval of the presented version.

