## Supplementary for Brenner for "Improved pathogen identification in sepsis or septic shock by clinical metagenomic sequencing"

**<u>This PDF file includes:</u>**

**Supplemental Methods**

Data collection

Next-Generation Sequencing data analyses

Web-based expert panel evaluations

**References**

**Supplemental Tables**

Supplemental Table 1

Supplemental Table 2

Supplemental Table 3

Supplemental Table 4

Supplemental Table 5

Supplemental Table 6

Supplemental Table 7

Supplemental Table 8

Supplemental Table 9a

Supplemental Table 9b

**Supplemental Figure Legends**

**Supplemental Figures**

Supplemental Figure 1

Supplemental Figure 2

Supplemental Figure 3

Supplemental Figure 4

#### Supplemental Methods

##### Data collection

Upon enrolment at sepsis onset the following baseline data were collected: demographics, date and time of hospital and ICU admission, admission source, major comorbid conditions (as assessed by the Charlson Comorbidity Index (CCI)) (1), immunosuppressive host factors (2), site of suspected or confirmed infection, antimicrobial course prior to study enrolment, surgery/procedures for suspected site of infection prior to enrolment, and Sequential Organ Failure Assessment (SOFA) Score. Clinical data collection at study inclusion (=Onset) as well as 72 hours afterwards (=72h) included, among other things, relevant laboratory data, antimicrobial/ antibiotic therapy, use of mechanical ventilation, vasoactive therapy, renal replacement therapy, surgical and other interventional procedures as well as radiological findings. The final outcome evaluation was performed at 28 days and included the following items: date of discharge, discharge destination and vital status at discharge. All data were recorded in standardized electronic case report forms (eCRF) using REDCap.

##### Next-Generation Sequencing data analyses

In brief, sequencing data were prepared as described before: about 30 million raw reads per sample were processed with the standard pipeline. All normalized non-human reads from blood plasma samples were assigned to the species level using Kraken 1 classification, based on the NCBI RefSeq database of microbial reference and completed genomes. A control cohort without any clinical indications of sepsis or infection was used for the calculation of quantitative expectation values for every species as background. The SIQ-score analysis involves a comparison of every clinical sample with the mean abundance of every detected species among the control cohort and a comparison with the predefined thresholds. It quantifies the significance of species detection relative to a control cohort of non-septic post-operative patients and is calculated as the deviation of normalized species abundance from baseline levels observed in the cohort, accounting for batch-specific positive and negative controls to remove technical noise. The control cohort used in this study improves the precision of the SIQ score calculation. According to modified pipeline, a species is considered for further analysis only if its normalized read count exceeds a baseline threshold of 3 (compared to a stricter threshold in earlier studies). The threshold for species detection was set to SIQ > 10 due to a plausible signal-to-noise ratio. Additionally, species are further filtered based on batch-specific controls and published bona fide contaminant lists (3). However, if SIQ scores for bona fide contaminants were > 500 species were called as relevant pathogens (**Supplemental Figure 1**). Calling of some exemplary species across all cohort samples based on the decision tree is shown in **Supplemental Figure 2**.

**Web-based expert panel evaluations**

Reports for expert panel evaluations were generated by compiling all clinical data from REDCap as well as NGS-based results. The REDCap database contains more than 800 clinical parameters for every patient, collected during their hospital stay. Data for all patients were downloaded from the REDCap database as a tab-delimited text table and underwent post-processing using scripts written in R statistical language, which includes a decision tree-based algorithm to convert table representation of the data to a human-readable text form. The patient report contains 4 different sections: description of a clinical case; clinical course and laboratory measurements; antibiotic treatment; NGS and microbiology section. Every patient report was manually curated by 3 individuals to avoid inconsistencies. An exemplary report is included in supplementary materials (**Supplemental Figure 3**). All presented patients' data were stored according to safety regulations and data compliance policies. Every questionnaire accompanying the reports contained three standard questions, aimed at the plausibility of the NGS-based results and their relevance for therapeutic intervention (**Supplemental Figure 4**).

### Supplemental Tables.

**Supplemental Table 1:** Inclusion and exclusion criteria of the *Next GeneSiS-Trial*.

| | |
|---|---|
| **Inclusion criteria** | |
| Age ≥18yr | |
| Informed consent | |
| Sepsis (with an onset ≤24h) | Patients with a life-threatening organ dysfunction caused by a dysregulated host response to a suspected or proven infection. Organ dysfunction can be identified as an acute change in total SOFA score ≥2 points consequent to the infection. Patients can also be promptly identified at the bedside with qSOFA, ie, alteration in mental status, systolic blood pressure ≤100mmHg, or respiratory rate ≥22/min. |
| *or* Septic shock (with an onset ≤24h) | Patients with septic shock can be identified with a clinical construct of sepsis with persisting hypotension requiring vasopressors to maintain mean arterial pressure (MAP) ≥65mmHg and having a serum lactate level >2mmol/L (18mg/dL) despite adequate volume resuscitation. |
| **Exclusion criteria** | |
| Age <18yr | |
| Refusal to give consent | |
| Patient will probably be discharged from the ICU within the first 72 hours following inclusion | |
| Palliative treatment intent | |
| Clinician is not committed to aggressive treatment | |
| Death is deemed imminent and inevitable | |
| Patients who had previously been included, but are readmitted to the ICU during the same hospitalization, will not be included a second time. | |
| Abbreviations: MAP, mean arterial pressure; ICU, intensive care unit; (q)SOFA, (quick) Sequential Organ Failure Assessment-Score | |

**Supplemental Table 2.** Antiinfective treatment regimens.

| **Antiinfective treatment** (up to 6 days following study inclusion) | |
|---|---|
| Average no. of antiinfectives used per patient | 2.96±1.75 |
| Total no. of antiinfectives in all patients | 1453 |
| - Antibiotics: | 1288 (88.6%) |
| • Penicillins | 407 (28.0%) |
| • Carbapenems | 278 (19.1%) |
| • Cephalosporins | 103 (7.1%) |
| • Fluorochinolones | 44 (3.0%) |
| • Macrolides | 66 (4.5%) |
| • Glycopeptides | 159 (10.9%) |
| • Oxazolidinones | 58 (4.0%) |
| • Nitroimidazoles | 58 (4.0%) |
| • Others | 115 (7.9%) |
| - Antifungals: | 131 (9.0%) |
| • Polyenes | 2 (0.1%) |
| • Azoles | 57 (3.9%) |
| • Echinocandins | 71 (4.9%) |
| • Others | 1 (0.1%) |
| - Virostatic agents: | 34 (2.3%) |
| • DNA-polymerase inhibitors | 32 (2,2%) |
| • Others | 2 (0.1%) |
| TDM-based guidance of antiinfective treatment | 263 (18.1%) |

Data are presented by number (%) or mean ± standard deviation.

Abbreviations: DNA, deoxyribonucleic acid, TDM, therapeutic drug monitoring

**Supplemental Table 3.** Microbiological samples.

| **Microbiological samples** (from 3 days prior to study inclusion up to 6 days following study inclusion) | |
|---|---|
| Average no. of microbiological samples per patient | 16.6±7.2 |
| Total no. of microbiological samples in all patients | 8167 |
| - Blood culture | 5536 (67.8%) |
| - Tracheal secretion | 493 (6.0%) |
| - Drainage fluid | 132 (1.6%) |
| - Swab samples from surgical site | 297 (3.6%) |
| - Wound swab | 147 (1.8%) |
| - Punctate | 65 (0.8%) |
| - Catheter tip | 140 (1.7%) |
| - Urine sample | 527 (6.5%) |
| - Cerebrospinal fluid | 23 (0.3%) |
| - Tissue sample | 53 (0.6%) |
| - Hygiene screening | 721 (8.8%) |
| - Stool sample | 30 (0.4%) |

Data are presented by number (%) or by mean ± standard deviation.

**Supplemental Table 4.** Species detection in NGS-/BC+ subgroup samples. The table summarizes cases where species were identified by blood culture (BC) and their corresponding detection status by next generation sequencing (NGS). Most species listed are common contaminants or commensals, highlighting the challenges of interpreting BC results. For NGS, species marked as contaminants or below the significance threshold were not called, reflecting the pipeline's ability to filter out low-confidence or background signals.

| Patient | NGS | BC |
|---|---|---|
| # | *Staphylococcus epidermidis* - detected, marked as a contaminant | *Staphylococcus epidermidis, Staphylococcus hominis* |
| # | *Propionibacterium acnes* - detected, marked as a contaminant | *Cutibacterium acnes* |
| # | *Cutibacterium acnes* detected - at OnSet - positive, at day 7 - marked as a contaminant | *Cutibacterium acnes, Streptokokkus intermdius* |
| # | *Staphylococcus epidermidis* - detected, below significance threshold | *Staphylococcus epidermidis* |
| # | not detected | *Staphylococcus epidermidis* |
| ***#*** | *Clostridium symbiosis* - not in the Kraken DB, used for the analysis; *Robinsoniella peoriensis* - not detected | ***Clostridium symbiosis, Robinsoniella peoriensis*** |
| # | *Enterobacter cloacae* - detected, below significance threshold | *Enterobacter cloacae* |
| ***#*** | *Klebsiella oxytoca, Klebsiella pneumoniae* - detected, below significance threshold | ***Klebsiella oxytoca, Klebsiella pneumoniae*** |
| # | *Staphylococcus epidermidis* detected - marked as a contaminant | *Staphylococcus epidermidis* |
| ***#*** | *Streptococcus pneumoniae* - at OnSet detected below significance threshold; 72h - detected | ***Streptococcus pneumoniae*** |
| # | not detected | *Staphylococcus epidermidis* |
| # | *Staphylococcus epidermidis* detected - marked as a contaminant; *Staphylococcus haemolyticus, Staphylococcus lugdunensis, Staphylococcus saprophyticus* - not detected | *Staphylococcus epidermidis, Staphylococcus haemolyticus, Staphylococcus lugdunensis, Staphylococcus saprophyticus* |
| # | not detected | *Staphylococcus epidermidis* |
| # | not detected | *Staphylococcus saccharlyticus* |
| # | *Staphylococcus epidermidis* detected - marked as a contaminant; *Staphylococcus capitis* - not detected | *Staphylococcus capitis, Staphylococcus epidermidis* |
| # | not detected | *Staphylococcus capitis, Staphylococcus hominis, Staphylococcus epidermidis* |

**Supplemental Table 5.** 2x2 contingency table for NGS vs. BC concordance.

| total | NGS + | NGS - | ∑ |
|---|---|---|---|
| BC + | 158 | 16 | 174 |
| BC - | 466 | 248 | 714 |
| ∑ | 624 | 264 | |

**Supplemental Table 6.** Percentage concordance at onset and day 3 between NGS vs. BC.

| | Onset | Day 3 | Total |
|---|---|---|---|
| matching BC/NGS | 47.6% | 43.4% | 45.7% |
| unmatching BC/NGS | 52.4% | 56.6% | 54.3% |

**Supplemental Table 7.** Statistical association between NGS vs. BC results.

| | |
|---|---|
| Chi-Square Statistic | 42.46 |
| p-value (Chi-Square) | 7.20E-11 |
| Degrees of Freedom | 1 |
| Odds Ratio (Fisher) | 5.26 |
| p-value (Fisher) | 1.36E-12 |

**Supplemental Table 8a.** Treatment recommendations and clinical outcome characteristics in a NGS(+) and BC(-) subgroup of septic patients.

| **Outcomes (at or within 28 days after study inclusion)** | **Treatment recommendation** | | | | | **p-value** |
|---|---|---|---|---|---|---|
| | **Maintenance (n=112)** | **Escalation (n=23)** | **De-escalation (n=56)** | **Combination (n=5)** | **Total (n=196)** | |
| Mortality | 17 (15.2%) | 6 (26.1%) | 13 (23.2%) | 1 (20.0%) | 37 (18.9%) | 0.480 |
| Length of ICU/IMC stay [days] | 11.7±8.4 | 12.7±8.7 | 15.4±9.3 | 13.8±9.6 | 12.9±8.8 | 0.087 |
| Rel. length of ICU/IMC stay [days] | 0.54±0.41 | 0.56±0.38 | 0.69±0.36 | 0.63±0.43 | 0.59±0.39 | 0.168 |
| Length of mechanical ventilation [days] | 5.8±7.3 | 8.0±9.2 | 9.6±9.8 | 6.2±6.5 | 7.2±8.4 | 0.050 |
| Rel. length of mechanical ventilation [days] | 0.32±0.40 | 0.36±0.42 | 0.44±0.41 | 0.25±0.22 | 0.35±0.41 | 0.264 |
| Length of antibiotic treatment [days] | 11.7±7.4 | 12.7±7.7 | 14.7±8.2 | 15.0±9.8 | 12.8±7.8 | 0.107 |
| Rel. length of antibiotic treatment [days] | 0.53±0.36 | 0.55±0.32 | 0.65±0.32 | 0.62±0.33 | 0.57±0.35 | 0.191 |
| Length of antifungal treatment [days] | 3.1±6.6 | 1.7±5.4 | 5.5±6.7 | 2.2±3.0 | 3.6±6.5 | 0.054 |
| Rel. length of antifungal treatment [days] | 0.13±0.28 | 0.06±0.2 | 0.24±0.30 | 0.14±0.24 | 0.16±0.28 | 0.051 |
| Length of antiviral treatment [days] | 0.6±2.2 | 0.8±2.7 | 2.6±5.8 | 0±0 | 1.2±3.7 | **0.006**** |
| Rel. length of antiviral treatment [days] | 0.02±0.08 | 0.03±0.10 | 0.11±0.25 | 0±0 | 0.05±0.15 | **0.002**** |
| Length of KRT [days] | 1.7±4.8 | 1.0±3.6 | 5.9±9.6 | 2.0±4.5 | 2.8±6.7 | **<0.001***** |
| Rel. length of KRT [days] | 0.09±0.22 | 0.06±0.16 | 0.24±0.37 | 0.18±0.41 | 0.13±0.28 | **0.003**** |

Relative data are related to the 28-day observation period (for patients who survived the 28-day observation period) or to the survival duration (for patients who died within the 28-day observation period). Data are presented by number (%) or mean ± standard deviation. Concerning symbolism and higher orders of significance: *p < 0.05, **p < 0.01, ***p < 0.001

Abbreviations: BC, blood culture; ICU, intensive care unit; IMC, intermediate care; KRT, kidney replacement therapy; NGS, next generation sequencing

**Supplemental Table 8b.** Treatment recommendations and absolute clinical outcome characteristics in a NGS(+) and BC(-) subgroup of surviving septic patients.

| **Absolute outcomes (at or within 28 days after study inclusion)** | **Treatment recommendation** | | | | | **p-value** |
|---|---|---|---|---|---|---|
| | **Maintenance (n=95)** | **Escalation (n=17)** | **De-escalation (n=43)** | **Combination (n=4)** | **Total (n=159)** | |
| Length of ICU/IMC stay [days] | 12.2±8.9 | 12.7±9.3 | 16.7±9.8 | 14.3±11.x | 13.5±9.3 | 0.077 |
| Length of mechanical ventilation [days] | 5.6±7.7 | 7.1±9.2 | 10.0±10.5 | 7.0±7,2 | 7.0±8.8 | 0.063 |
| Length of antibiotic treatment [days] | 12.2±7.7 | 13.1±8.3 | 15.9±8.4 | 16.8±10.0 | 13.4±8.1 | 0.080 |
| Length of antifungal treatment [days] | 3.3±7.0 | 1.5±6.1 | 5.7±7.0 | 1.2±2.5 | 3.7±6.9 | 0.097 |
| Length of antiviral treatment [days] | 0.07±2.4 | 1.1±3.1 | 2.9±6.2 | 0±0 | 1.3±4.0 | **0.015*** |
| Length of KRT [days] | 1.7±5.0 | 0.06±0.24 | 6.4±10.2 | 0±0 | 2.7±6.9 | **<0.001***** |

Data are presented by number (%) or mean ± standard deviation. Concerning symbolism and higher orders of significance: *p < 0.05, **p < 0.01, ***p < 0.001

Abbreviations: BC, blood culture; ICU, intensive care unit; IMC, intermediate care; KRT, kidney replacement therapy; NGS, next generation sequencing

**Supplemental Figure legends.**

**Supplemental Figure 1.** Decision tree for calling relevant pathogenic species in patient samples by NGS data.

The flowchart illustrates the decision-making process for species identification from next-generation sequencing data. At each decision point (box), conditions are evaluated, leading to actions (diamonds) or terminal outcomes (circles). Arrows indicate the logical flow, with red text highlighting calling decisions.

**Supplemental Figure 2.** Normalized species count distributions across samples for selected microorganisms.

This figure shows the distributions of normalized species counts for four selected species (*Escherichia coli, Bacteroides fragilis, Enterococcus faecium, and Human herpesvirus 5*). Each row corresponds to one species, with two panels per species. (a) Full-scale normalized species count distributions across all samples. (b) Zoomed-in view of normalized species counts with the y-axis limited to 50 to highlight low-abundance features. Points are color-coded to represent different detection categories: red indicates called features (detected and significant), orange represents detected but not significant features (not called), and green denotes features that were not detected; grey dots represent biological control (POP) and dark yellow dots represent technical control samples. Data are plotted for all sequenced samples, regardless of sampling day. Y-axis values shows normalized species abundance.

**Supplemental Figure 3.** Exemplary patient report.

A large number of pathogens has been identified by next-generation sequencing (NGS) at sepsis onset (day 0), as well as at 3 days after sepsis onset. The blood culture test was negative for both time points. The analysis of a surgical swap didn't show any clinically relevant species, as stated by the medical personal (indicated by *). Species which were found in significant quantities compared to the other day are shown in bold type.

<u>Abbreviations:</u> CRP, c-reactive protein; ICU, intensive care unit; IMC, intermediate care unit; NA, not applicable; PCT, procalcitonin; SOFA, sequential organ failure assessment

**Supplemental Figure 4.** Online survey questions.

Questions q1 and q2 are obligatory to answer. Question q3 is connected to question q2 and is demonstrated to experts only if question q2 is answered with “No”.

<u>Abbreviations:</u> NGS, next generation sequencing

**Supplemental Figures.**

**Supplemental Figure 1.** Decision tree for calling relevant pathogenic species in patient samples. by NGS data.

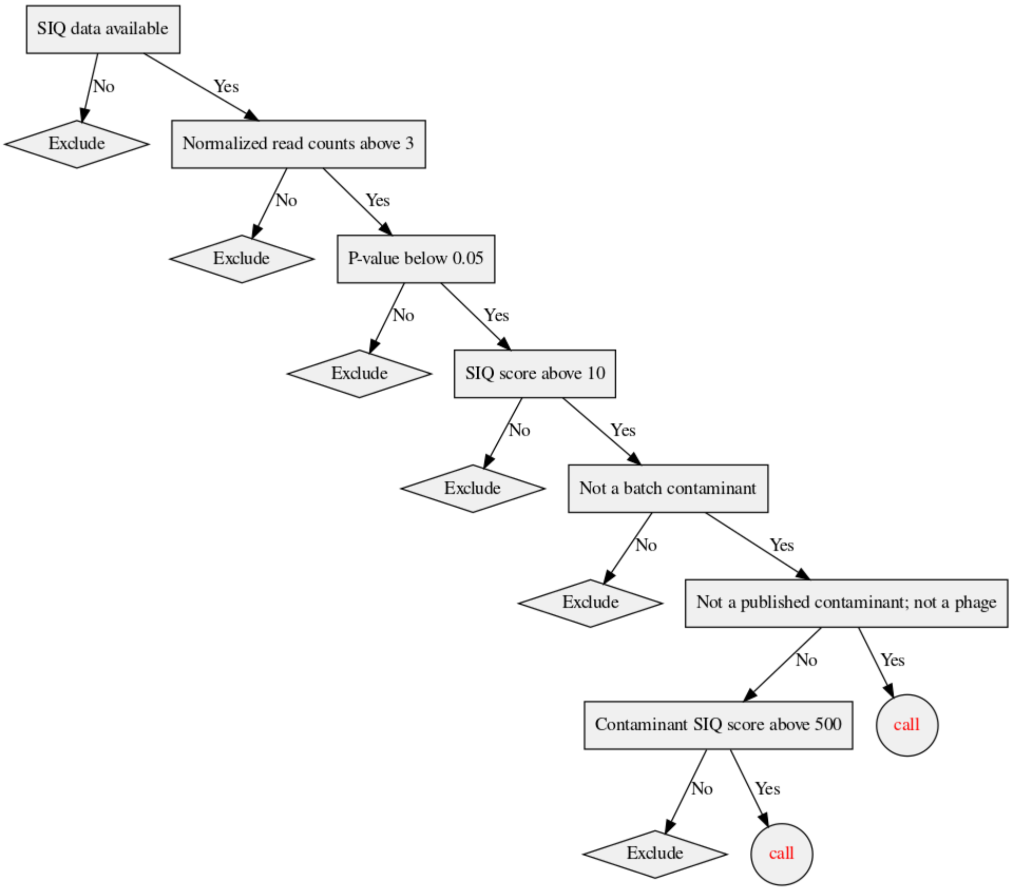

**Supplemental Figure 2.** Normalized species count distributions across samples for selected microorganisms.

a. *Escherichia coli*

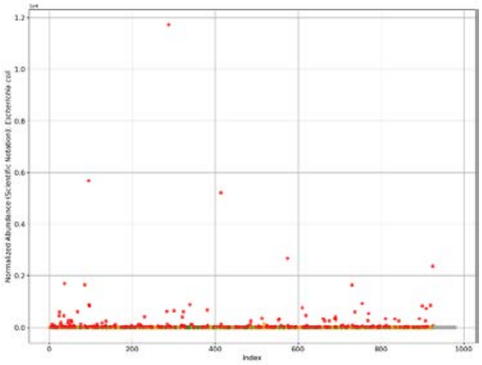

b.

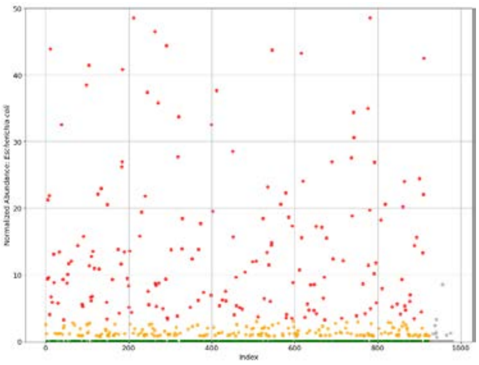

c. *Bacteroides fragilis*

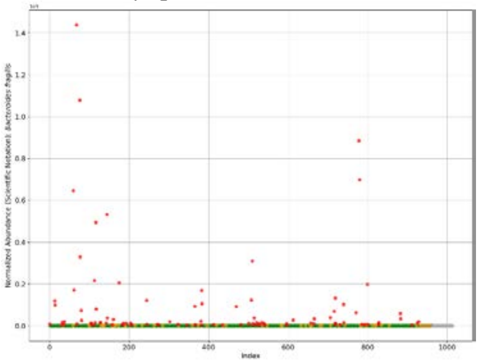

d.

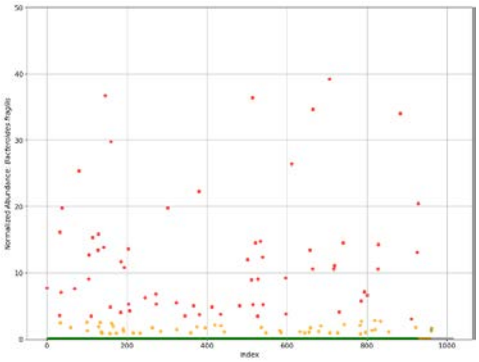

e. *Enterococcus faecium*

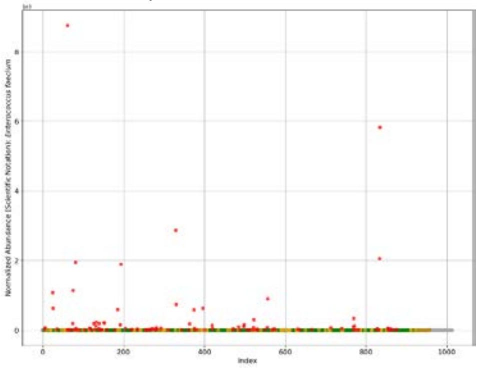

f.

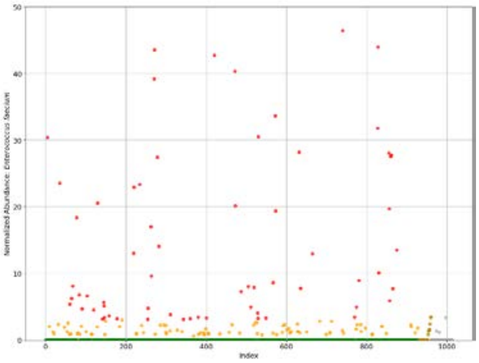

i. *Human herpesvirus 5*

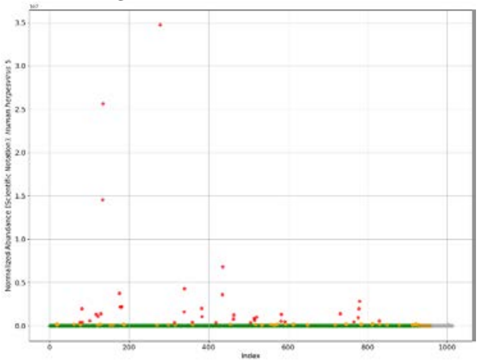

j.

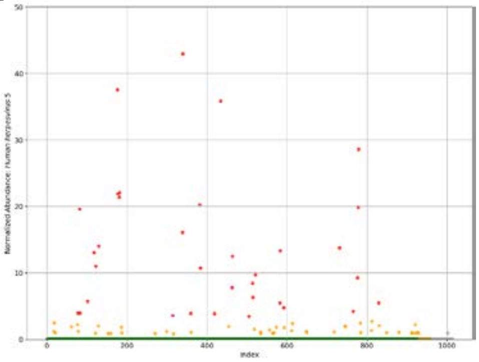

**Supplemental Figure 3.** Exemplary patient report.

#### Patient XYZ

**Anamnesis**
The patient is between 70-80 years old and a surgical emergency who was on a normal ward before the admission to the ICU.

**Focus of primary infection**
On day 0 (sepsis onset), clinically suspected ambulant sepsis with urogenital localisation and septic shock are observed.
On day 3 after sepsis onset, clinically suspected ambulant sepsis with intra-abdominal localisation and septic shock are observed.
One day after sepsis onset a surgical focal decontamination was performed with the description „exploratory laprotomy, bride lysis, intestinal decompression".

**Outcome**
The duration of treatment in an ICU/IMC was 5 days. The study was terminated prematurely on day 4 due to patient death. The patient data collected is complete and verified.

#### Clinical course and laboratory measurments

| | Day 0 | Day 3 |
|---|---|---|
| SOFA | 8 | 12 |
| lactate | 2 mmol/l | 2.9 mmol/l |
| CRP | 69 - 103 mg/l | 72.3 mg/l |
| PCT | 0.23 - 1.12 µg/l | 5.41 - 6.14 ng/ml |
| leukocytes | 5.8 - 10.2 gpt/l | 4.5 gpt/l |

#### Anti-infectives

| | vor Onset | Tag 0 | Tag 1 | Tag 2 | Tag 3 | nach Tag 3 |
|---|---|---|---|---|---|---|
| Aminopenicillins +ß-lactamase inhibitor | | | | | | |

#### NGS & Microbiology

| | Day 0 | Day 3 | Other time points |
|---|---|---|---|
| NGS | ***Bacteroides fragilis***, *Akkermansia muciniphila*, ***Proteus mirabilis***, *Parabacteroides distasonis*, ***Escherichia coli***, ***Bacteroides vulgatus***, ***Bacteroides thetaiotaomicron*** | *Akkermansia muciniphila, Bacteroides fragilis, Parabacteroides distasonis, Escherichia coli*, ***Faecalibacterium prausnitzii***, *Bacteroides vulgatus*, ***Eubacterium rectale*** | NA |
| Blutkultur aerob | Negativ | Negativ | NA |
| Blutkultur anaerob | Negativ | Negativ | NA |
| *Andere Proben* | *Negativ* | *NA* | *Staphylokokkus epidermis*, Staphylokokkus haemolyticus*, Staphylokokkus saprophyticus*, Staphylokokkus lugdunensis* (OP-Abstrich)* |

** - Klinisch nicht relevanter Nachweis*

**Supplemental Figure 4.** Online survey questions.

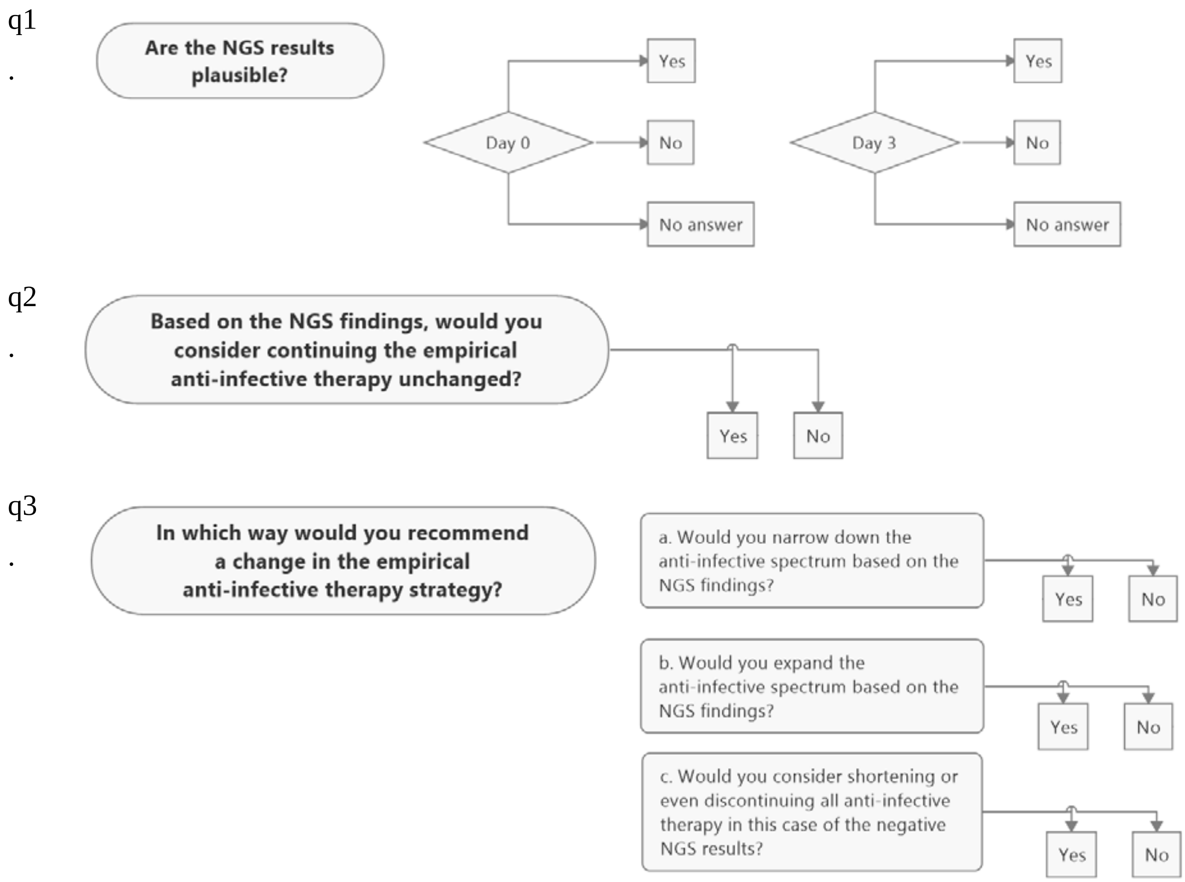
